# Appropriateness of video call consultations in a Dutch primary care setting

**DOI:** 10.1101/2022.12.15.22283509

**Authors:** Daniel Lindh, Eric Luiten

## Abstract

The pressure on healthcare is increasing worldwide. The trend toward digitalization in recent decades and the emergence of modern technologies have enabled remote treatment and offered patients new ways to interact with physicians. Despite numerous studies showing promising results in the use of video calls for primary care consultations, some survey studies report lingering doubts about the effectiveness of this method among general practitioners (GPs) and patients. Because the organization of health care varies widely across countries, we set out to investigate the appropriateness of video calling in a Dutch health care setting. Unlike many survey studies, we collected real-life data where GPs evaluated the video call directly after the consultation, allowing a more realistic assessment of perceived quality. We find that video calls are appropriate 84.2% (95% CI 80.3%-87.8%) of the time for consultations with patients of moderate to low urgency. Importantly, the appropriateness of video calls decreased with increasing urgency. These results build on previous studies and confirm the positive use cases of video calls focused on primary care in the Netherlands.

## Introduction

The pressure on health care is increasing worldwide. A general trend is that Western countries are dealing with an ageing population and thus with more chronically ill and multimorbid patients (Kringos et al., 2015), who often require frequent consultations and complex care (Sondergaard et al., 2015). Specifically in the Netherlands, general practitioners (GPs) play a significant role in access to health care where they directly solve about 90% of all health problems and complaints of their population (Cardol et al., 2004). In addition, the Netherlands has the highest average number of patients per GP compared to other countries where registration by name is common (Schäfer et al., 2016). This makes the Netherlands one of the countries where new innovations and solutions are most needed.

The trend in recent decades toward digitalization and the emergence of modern technologies has enabled remote treatment and offered patients new opportunities to interact with physicians.

Previous studies have reported that video calls, in comparison to telephone and face-to-face consultations, have a positive impact on the timeliness, availability, and effectiveness of health care (Carrillo de Albornoz et al., 2021; Sandbrink et al., 2021; Thiyagarajan et al., 2020; Turner et al., 2022). Furthermore, survey studies indicate that patients’ experiences with telemedicine are positive overall where patients report that it is easy to use, has a low cost, and reduces travel time (Sandbrink et al., 2021). In comparison to phone calls, videocalls allow GPs to observe the patient, receive nonverbal signals, and perform a limited physical examination making it a more attractive tool than phone calls. In fact, studies have shown that GPs who conduct a video consultation make more correct treatment decisions compared to those who conduct only telephone consultations (Capampangan et al., 2009; Handschu et al., 2008; Rush et al., 2018), as well as leading to reduced readmission rates (Rush et al., 2018). During the COVID-19 pandemic, patient use of telemedicine increased substantially (Johnsen et al., 2021), allowing patients to see their physician without risking infection. In the Netherlands, physical consultations decreased from 78% to 16% during the pandemic, while video consultations increased from 3% to 20% of all interactions between GP and patients (Sandbrink et al., 2021). This trend has been welcomed by policymakers around the world seeking to alleviate pressure on the overburdened healthcare system (*Health Canada’s Approach to Digital Health Technologies*, 2018; *NHS Long Term Plan*, 2022).

However, some studies suggest that skepticism persists among both GPs and patients, who still prefer in-person visits (Randhawa et al., 2018; Thiyagarajan et al., 2020), although meta-studies have found that consultations conducted via videocalls were as effective as in-person visits in treating adults (Carrillo de Albornoz et al., 2021). While many studies have examined the feasibility and attitudes toward video calls, there are few studies specific to the Dutch health care system. Moreover, many studies are based on surveys sent to patients and GPs, which are designed to measure GPs and patients’ expectations rather than direct experiences. Here, we fill this knowledge gap by surveying Dutch GPs directly after a videocall consultation to assess the appropriateness of the videocall compared with a face-to-face consultation. We aimed to build on previous studies that have investigated the appropriateness of different complaints (Brunett et al., 2015), but include a broader range of complaints. To anticipate: We found that video calls were appropriate for patient complaints about 84.2% of the time, with the most and least appropriate categories being general & skin and bodily injury, respectively. These results can guide various stakeholders, such as patients and primary care physicians, in making informed decisions when shaping Dutch healthcare.

## Methods

In this study, data on GPs’ experiences with video call consultations were collected between 2021 and 2022 at a general practice clinic in Amsterdam. A total of 361 Dutch-speaking patients who were not in a life-threatening situation and 13 primary care physicians (women = 7) with at least 3 years of experience recruited by Quin to perform video calls participated. None of the GPs were the patient’s own GP, but a locum GP working at a central location. The working hours of the video consultants were the same as those of the regular GPs. Informed consent was obtained from all GPs. Patients gave consent to share their digital symptom checker result with their GP. We did not collect any other demographic information, so no further patient consent was required according to the Centrale Commissie Mensgebonden Onderzoek (CCMO). Personal data were handled in a pseudo-anonymous manner and in accordance with the EU General Data Protection Regulation (GDPR) and the Dutch Act implementing the General Data Protection Regulation (Uitvoeringswet AVG). Because the study did not fall under the Medical Research Involving Human Subjects Act (WMO), we did not require prior ethical approval by the Medical Research Ethics Committee (MREC) or the CCMO.

### Procedure

Before booking a video consultation, patients completed an assessment using a digital symptom checker (developed by Quin B.V., The Netherlands) that ranked the urgency level of their complaints from U1 - “Call 122 or the practice’s emergency number GP “ - to U5b - “No appointment needed.” (See *Categorizations* below). Only patients with an urgency level of U3, U4, and U5a were eligible for this study, in which they were offered to complete their consultation via video call with a locum GP. After completing the consultation, the GP was asked to complete questionnaires on the feasibility and satisfaction with the video call. After completing the questionnaires, the GP was also asked to assign a level of urgency to the patient’s health complaints. See Figure 1A for a pictorial representation of the workflow. Data on health complaints, wait time for a video call, and length of consultation were extracted from the digital symptom checker and the Quin platform.

**Figure 1.**
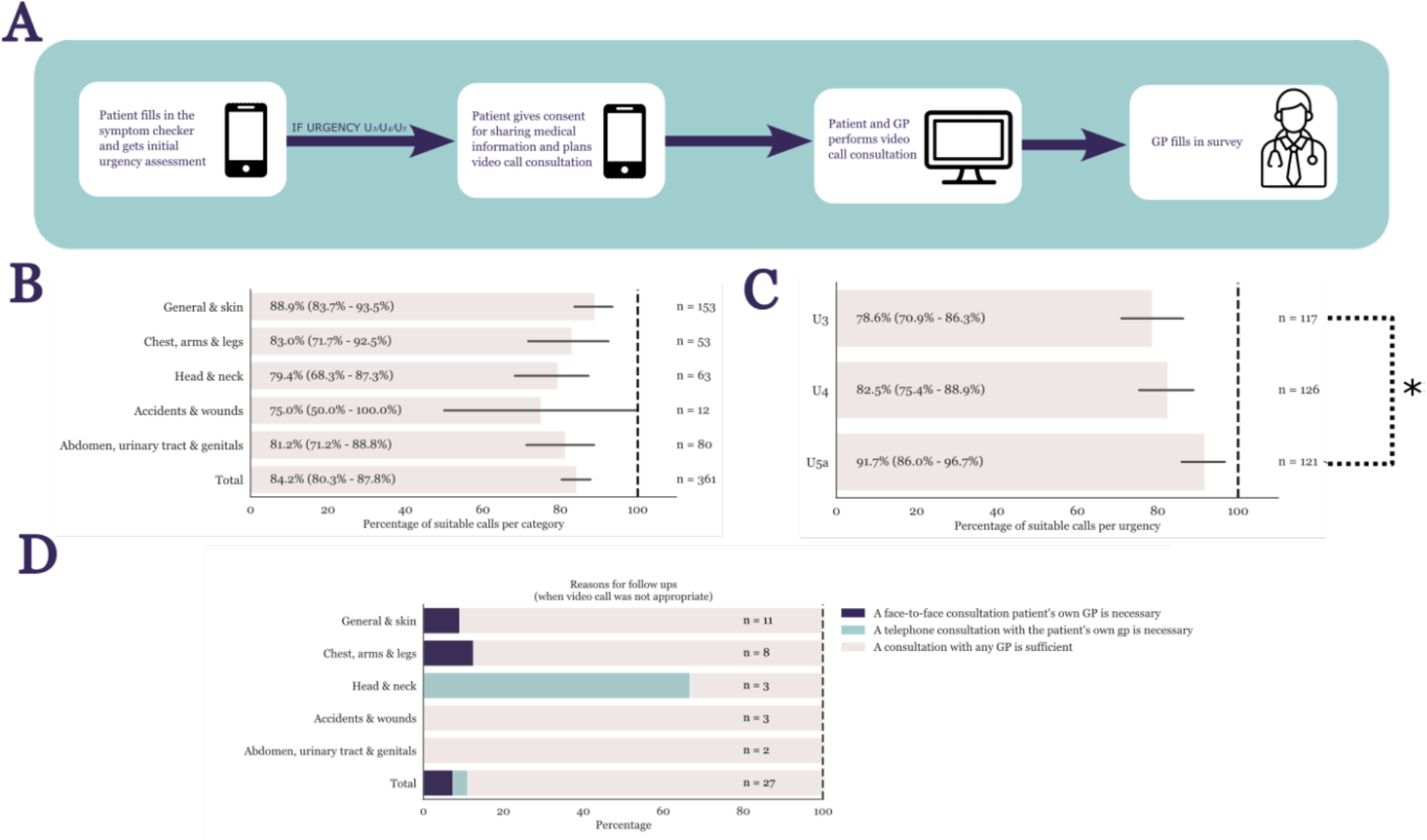
Experimental setup and main results. A) Pictorial representation of workflow. B) Percentage of cases when a video call was suitable for the patient complaint for each high-level medical category. Accidents and wounds were the least suitable category while general and skin-related issues were highly suitable for video calls. However, no statistical difference was found between categories. C) Percentage of cases when a video call was suitable for the patient complaint for each urgency-level. An ANOVA revealed a significant difference between urgency levels (See Methods – Categorization) in suitability of video call. D) Suitable follow-ups (when necessary) were mostly sufficiently covered with any type of GP and not the patient’s own GP. The low total of responses is due to it not being a mandatory question for the GP. Black lines as well as values inside parenthesis on the bar plots indicate 95% confidence interval. * = p < 0.05, ** = p < 0.01.

### Categorizations

In total, we have obtained 57 distinct types of chief complaints. However, for simplicity, we categorized them into five categories used by the Nederlandse Triage Standaard (NTS): ‘General & Skin’, ‘Chest, Arms & Legs’, ‘Head & Neck’, ‘Accidents & Wounds’, ‘Abdomen, Urinary Tract & Genitals’. We further categorized the urgency levels according to NTS: U1: Call an ambulance, U2: Contact your doctor immediately, U3: Contact GP within a few hours, U4: Make an appointment within 24 hours, U5a: Make an appointment within a week, U5b: self-care at home. However, U1, U2, and U5b cases were excluded from the study.

### Statistical analysis

The results of the feasibility and satisfaction questionnaire and the symptom checker and Quin platform data were analyzed using Python (version 3.7) and JASP (JASP Team (2022), JASP (Version 0.16.3)), see [ADDRESS TO CODE/DATA REPOSITORY] for code and data. The data on video consultations (urgency levels and complaints) and the results of the two questionnaires were described using standard descriptive statistics (frequencies and percentages).

## Results

The present study aimed to evaluate the suitability of video calls for consultation between patients and GPs in a Dutch health care setting. We collected data from 361 consultations in a period between 2021-2022. A Shapiro-Wilk test for normality showed a significant deviation from normality (W=0.885, p < 0.001) for consultation durations, therefore Wilcoxon signed rank test and Kruskal-Wallis tests were chosen for further comparisons. The mean duration of each consultation was significantly shorter than the previously reported face-to-face consultation of 10.2 minutes (Deveugele et al., 2002), with mean = 7.62 minutes (95% CIs, 7.15-8.09 minutes), SD = 4.56, median = 7 minutes. Wilcoxon signed rank T(363) = -9.942, Z=-52.278, p <.001. A Kruskal-Wallis test revealed a significant difference in consultation time between urgency levels, with U3 (M=8.59 minutes, SD =4.6, N=117), U4 (M=6.5, SD =4.7, N=121), and U5a (M=8.55, SD =6.1, N=121), H(2) = 17.144, p < 0.001. See *Methods – Categorizations* for definitions of urgency levels.

Overall, primary care physicians rated the video consultation as appropriate for the patient’s complaint in 84.2% (95% CI 80.3%-87.8%, N=361) of cases (see Figure 1). When broken down by our complaint categories, video call was considered appropriate: General & Skin (M=88.9% of the time, 95% CI 83.7%-93.5%, N=153); Chest, Arms, and Legs (M=83%, 95% CI 71.7%-92.5%, N=53); Head and Neck (M=79.4%, 95% CI 68.3%-87.5%, N=63); accidents and wounds (M=75%, 95% CI 50%-100%, N=12); abdomen, urinary tract, and genitals (M=81.2%, 95% CI 80.3%-87.8%, N=80). A Kruskal-Wallis test showed no significant difference between the categories, H(4) = 4.824 p = 0.304. If it was indicated that a video call was not appropriate for the complaint, GP had the option to indicate why this was not the case (although this was not mandatory). A total of 27 responses were collected, with a follow-up physical consultation with any GP being sufficient in most cases (81.4%), followed by “an in-person consultation with one’s GP is necessary” (11.1%) and “a phone call with the patient’s own GP is necessary” (7.4%). Grouped by urgency, we also found that U3 (M=78.6%, 95% CI 70.9%-86.3%, N=117); U4 (M=82.5%, 95% CI 75.4%-88.9%, N=126); U5a (M=91.5%, 95% CI 86.4%-95.8%, N=118); X^2^(2, N=361) = 8.722, p = 0.012, and the predictor variables U3 and U5a contributed significantly to the logistic regression model, with U3 B = 1.303, SE = 0.226, Wald = 33.371, p < 0.001; U4 B = 0.25, SE = 0.325, Wald = 0.592, p = 0.442; U5a B = 1.104, SE = 0.4, Wald = 7.624, p = 0.006.

## Discussion

In the Netherlands, the need for solutions to mitigate the pressures in primary healthcare is growing due to population growth and urbanization (*Statistics Netherlands* (*Cbs*.*Nl)*, 2022). In addition, the demand for better access to care is increasing due to the aging population and the rise in chronic diseases such as diabetes, cancer, or heart disease (*Statistics Netherlands* (*Cbs.Nl*), 2022). One such solution is the introduction of video consultations as opposed to in-person consultations. Video consultations have many advantages, such as no travel for the patient, better accessibility for people living in remote areas, and no risk of contracting infectious viruses in the clinic (Sandbrink et al., 2021). Compared to phone calls, video call consultations allow for visual inspection as well as more dynamic interaction (Donaghy et al., 2019; McKinstry et al., 2009). Although many previous studies have shown that video calls are comparable to face-to-face consultations in many cases (Dixon & Stahl, 2009; Thiyagarajan et al., 2020) some survey studies still indicate that both patients and GPs are doubtful to the use of video calls (Randhawa et al., 2018; Thiyagarajan et al., 2020). Here we asked GPs to rate the appropriateness of a video call immediately after the consultation so that we can see how video calls are performing as a consultation medium in practice.

Previous studies on video calls in primary care have shown that they are comparable to face-to-face consultations (Carrillo de Albornoz et al., 2021). However, face-to-face consultations are still preferred by patients and primary care physicians (Randhawa et al., 2018; Thiyagarajan et al., 2020). One of the most common reasons for the reluctance among GPs is the fear that there will be a loss of information because video calls can be less personal (Randhawa et al., 2018), and loss of time due to technical issues (Sandbrink et al., 2021). However, before the 2020 pandemic, 19% of consultations were done over the telephone (Sandbrink et al., 2021). A medium that has clear disadvantages against video calls, such as the inability to notice nonverbal signals and ability to perform visual examination. Although face-to-face consultations are preferred by GPs, researchers and GPs have also pointed out some disadvantages, such as travel time, travel time, cost, environmental impact, and risk of infection on public transport (Sandbrink et al., 2021). Disadvantages also included ways to mitigate the risk of infection, such as social distancing, hygiene measures, ventilation, and limiting the number of people in the waiting room. These are factors that are still relevant outside of the 2019 COVID pandemic (Shaw, 2019). Time is lost in picking up the client, putting on and taking off the gown, and discussing incidents. Information cannot be entered into the system until after the consultation: It is not always possible to type during the consultation. Scheduling consultations can be difficult when consultations take place in various locations. An on-site consultation room can be noisy. Sometimes the doctor is alone there, which can become a problem if (physical) aggression occurs.

Notwithstanding some reservations on the part of GPs and patients, most studies report positive outcomes from video calling. In a study of over 700 clinicians, Glaser et al. (2009) found that 88.2% (650/737) felt that a video call consultation had improved the patient’s prognosis, and 89.4% of clinicians (652/729) felt that clinical decision making was improved by video calls. Hammersley et al. (2019) reported that video calls were shorter than in-person consultations, but the content of the consultation was different because medical problems were discussed in more detail during in-person consultations. However, this study was a nonrandomized and pseudo experimental study, so a likely explanation for these results is differences in demographics and types of complaints. In a randomized control trial (RCT), Dixon and Stahl (2009) found no difference in patients’ perceptions of counseling between video calls and face-to-face counseling. However, GPs were less satisfied with video calls when new treatments were initiated, and they were less satisfied with their ability to order appropriate laboratory tests. These results were determined using a Likert scale of 1-5. Although statistically significant, the significance of a difference between 4.5 and 4.2 (out of 5) may be considered negligible in practice. Aside from this study, which is one of the few exceptions, there is a dearth of RCTs throughout the literature examining the difference between video calls and face-to-face consultations in primary care. This is a critical issue for appropriate scientific evaluation, as convenient sampling often results in differences in the health issues and demographic characteristics studied. Patients who consent to video consultations tend to be younger, healthier, and have less severe health problems. Again, we did not examine this issue in the current study. Instead, we focused on the extent to which video calls are appropriate for health problems encountered in a real Dutch primary care setting. This is an important first step. However, for video calling to help alleviate pressure on primary care, we need further studies that examine medical cases in the elderly population with more complex health problems, as this population is the most time-consuming and represents the target group for future technological innovations.

Similar to many other studies, we find that video calls are overall appropriate for most types of health complaints. It is important to note that in our study, patients self-selected to receive counseling via video call. This means that, overall, patients can correctly decide for themselves when to make a video call, a question many doctors are concerned about. We also find that video calls take significantly less time than the average face-to-face consultation in the Netherlands. This is consistent with the study by Dixon and Stahl (2009) and many other studies (e.g., Hammersley et al., 2019). There are two main arguments that can be made against video calls being shorter than face-to-face consultations. One could argue that patients who opt for a video consultation (1) may be younger and (2) have less serious health problems. However, because Dixon and Stahls’ study was an RCT study, and patients were randomly assigned to either a video consultation or an in-person consultation, differences in demographic and severity of illness cannot fully explain this effect. In addition, we find that although there is a significant difference in consultation time between urgency levels derived from our symptom checker, the urgency level that takes the least time is U4 (an average of 6.5 minutes versus 8.5 minutes for U3 and U5a). This means that consultation time does not increase with more severe health complications, which argues against the second caveat above. We also found that the triage levels of Quin’s own symptom checker are useful in predicting whether a consultation is appropriate for a video call, with a video call becoming increasingly less appropriate for more urgent health complaints. This symptom assessment tool was developed in-house (Quin has since the study upgraded their symptom checker to a higher performing one), and there is no reason to believe that it is superior to other symptom checkers on the market. This is important because it means that modern symptom checkers, some of which have similar accuracy in triaging as GPs (such as the Ada symptom checker; Gilbert et al., 2020), can be used to predict whether a consultation is appropriate for a video call or whether it is better done in person.

Given the many benefits that video calls bring, such as environmental friendliness, time savings, reduced risk of infectious disease, and accessibility (Carrillo de Albornoz et al., 2021; Thiyagarajan et al., 2020; Turner et al., 2022), it is imperative that we understand whether video calls are useful in practice. Here we show that, overall, video calls are appropriate for most types of conditions. Although we show numerical differences among our 5 categories of health complaints, we find no statistically significant difference. This could be due to our small sample size for some of the categories. We improved the methodology of other studies that rely on sending questionnaires or conducting interviews with GPs. We asked GPs to rate the consultation immediately after its completion, which allows the GP to evaluate the quality on a case-by-case basis. Considering our results in the context of the rest of the literature, video consultations are safe and appropriate for most health topics in a Dutch health care setting, and they tend to be shorter than face-to-face consultations. Considering that 84% of the Dutch population owns a smartphone that can do video calls (*Eurostat. Individuals Used a Mobile Phone (or Smart Phone) to Access the Internet*, 2020), it is a feasible implementation in many clinical practices. Future studies should focus on randomized trials to avoid issues related to convenience sampling. In addition, studies should examine how valuable video consultations can be for older people, taking into account both technological and health differences compared with younger generations.

## Data Availability

All data produced in the present study are available upon reasonable request to the authors

